# Interrater reliability of Tremor Characterization

**DOI:** 10.64898/2026.09.21.26363551

**Authors:** Prajakta Joshi, Yuri Ferreira Felloni Borges, Sanjay Pandey, Marie Vidailhet, Victor Fung, Mark Hallett, Hyder A. Jinnah, Aasef G. Shaikh, Alfonso Fasano

## Abstract

**Background:** Tremor is a repetitive, oscillatory movement with multiple subtypes requiring accurate characterization for diagnosis and treatment. While frequency, amplitude, and context are routinely used to describe tremor, recent recommendations have emphasized oscillation shape and regularity. However, these descriptors lack strict operational definitions, and differences in how clinicians interpret them introduce inconsistency into tremor classification.

**Objective:** We quantified inter-rater reliability for commonly used tremor descriptors and determined which objectively measured signal parameters: peak amplitude, peak frequency, and frequency variability best explained inconsistency in rater assessments.

**Methods:** We developed the Tremor Waveform Morphology Questionnaire (TWMQ) and obtained standardized video recordings of 34 individuals with upper-extremity tremor performing rest, postural, and kinetic tasks, with simultaneous kinematic recordings using an inertial sensor. Movement disorder experts rated tremor presence, shape, symmetry, regularity, rhythmicity, and diagnosis. We assessed inter-rater reliability, cross-feature dependencies, and modeled the influence of objective signal parameters on each subjective rating.

**Results:** Tremor presence and symmetry showed high agreement, whereas jerkiness, rhythmicity, amplitude stability, and clinical diagnosis showed lower reliability. Modeling revealed that peak amplitude primarily influenced judgments of tremor presence, jerkiness, and diagnosis, while frequency variability drove ratings of rhythmicity and amplitude stability.

**Conclusions:** Morphology-based descriptors remain subjective, but disagreement systematically reflects measurable signal properties. Objective definitions based on peak amplitude and frequency variability may improve the reliability and reproducibility of tremor characterization.

## Introduction

The term “tremor” often serves as an umbrella for involuntary rhythmic movements consisting multiple subtypes, each with distinct pathophysiological origins, making accurate characterization necessary for determining the appropriate diagnosis and treatment (1–8). Tremor can be present in isolation but also is a common feature of other movement disorders, including Parkinson’s disease, dystonia, and cerebellar dysfunction(6,9). In routine practice, clinicians describe tremor using features such as frequency (high vs. low), amplitude (high vs. low), distribution (body location and laterality), temporal pattern (consistent vs. intermittently present), and activation (rest vs. action) (10,11). Beyond these core descriptors, additional cycle shape-based descriptors such as regular vs. irregular amplitude, regular vs. irregular frequency, and jerky vs. sinusoidal waveform are sometimes used to infer underlying pathophysiology (8,11,12).

In the clinic, these parameters are rated during the physical exam based on the physician’s judgment, which can introduce subjective bias into the measures (4,11,13). Beyond quantitative assessment, bias may also arise from the lack of strict definitions, leaving key terms open to interpretation. Shape-based parameters lack operational guidelines, leaving evaluations to individual discretion (8,11,12). To better understand the rater decisions, we evaluated inter-rater reliability and examined whether rating one feature influenced ratings of others, underscoring the need for stronger guidelines, explicit definitions, and ultimately an objective tool to determine these parameters.

## Methods

### Participants and Ethical Approval

This study was performed in accordance with the Declaration of Helsinki and approved by the University of Toronto Institutional Review Board. All participants provided written informed consent prior to enrollment. Thirty-four adult patients (17 male, 17 female; mean age 65 ± 13 years) with a wide range of upper-extremity tremor severity were recruited. Diagnoses included essential tremor (ET; n = 18), Parkinson’s disease (PD; n = 6), dystonia with tremor (DT; n = 4), and other tremor etiologies (n = 6), including physiological tremor, drug-induced tremor, orthostatic tremor, functional tremor, and tardive tremor. The mean disease duration was 17.7 ± 14.8 years. Participants may have been receiving tremor-related medications at the time of examination. Videotaped recordings were obtained from individuals while performing standardized postures and tasks: “rest” (arms resting on the armchair), two types of “postural” tremors (outstretched position: arms extended at shoulder height, and “wing” position: forearms folded toward the chest with elbows lifted outward), and “kinetic” tremor (finger-to-nose).

### Ratings Acquisition

We developed a Tremor Waveform Morphology Questionnaire (TWMQ) to rate: (1) Presence of Tremor (Present/Absent); (2) Symmetry (Symmetric/Asymmetric/Unilateral); (3) Jerkiness (Absent/Present); (4) Amplitude Stability (Constant/Varying); (5) Rhythmicity (Regular/Irregular); (6) Frequency (Low/Mid/High); and (7) Diagnosis (ET/PD/DT/Other/No Tremor). For each participant, raters reviewed all posture or task segments and then selected the associated phenomenology on TWMQ. Ratings were performed by movement-disorders experts who routinely evaluate these features in clinical practice; all raters were blinded to diagnosis. Four movement disorder neurologists were recruited as raters for this reliability study. For the diagnosis item, the clinical diagnosis provided by the treating neurologist was also included and treated as a fifth rater. Raters assessed each tremor type for each participant individually; ratings were then collated for analysis.

### Assessment of Interrater Reliability

Interrater reliability looks at how consistently different raters rated the same item. Since there are more than two raters, and some missing data points, we selected Gwet’s AC1 (14) as the primary index of interrater reliability for nominal TWMQ items (Presence, Symmetry, Jerkiness, Amplitude Stability, Rhythmicity), and Gwet’s AC2 (quadratic weights) for the ordinal frequency item. We report AC2, 95% CIs (analytic/cluster bootstrap resampling by target), along with class balance and percent agreement.

### Combined Cross-feature Comparisons

To assess whether classification of one feature influenced judgments on another, we conducted cross-feature analyses for three prespecified pairs: (1) Amplitude Stability × Jerkiness, (2) Amplitude Stability × Rhythmicity, and (3) Rhythmicity × Jerkiness with ratings pooled across all raters. For each pair, we summarized contingency matrices and reported the corresponding conditional probabilities. For inference on cross-feature association, we applied χ² tests of independence (binary × binary) and reported effect sizes using φ (phi).

### Feature Dependence of Clinician Labels

To evaluate which signal features best explain clinician-assigned tremor labels, a permutation-based feature importance analysis was performed using an ensemble classifier. Six tremor qualities were assessed: presence of jerkiness, amplitude consistency, rhythmicity, overall tremor presence, perceived tremor frequency, and clinical diagnosis. The first four were binary labels (e.g., Present/Absent) rated by four independent neurologists. Tremor frequency was treated as a three-class ordinal label (Low/Mid/High) also rated by four neurologists. Clinical diagnosis was treated as a five-class nominal label (ET/PD/DT/Other/No Tremor) rated by five independent neurologists.

Tremor was objectively recorded for 30 seconds using an Xsens DOT inertial measurement unit (IMU; Movella Inc., Enschede, The Netherlands), which was secured to the dorsal aspect of the hand using a strap and positioned consistently across all recordings. Three quantitative signal features were extracted from each recording during three motor tasks (rest, postural wing-holding, and kinetic): frequency variability (FreqVar: *Defined as the bandwidth containing the central 66% of the signal’s spectral energy*), peak tremor frequency (peakF, defined as the frequency corresponding to the maximum spectral power), and peak tremor amplitude (peakAmp, defined as the maximum spectral power within the tremor frequency range). Since each recording was rated by multiple raters independently, feature vectors were replicated to align with each rater’s label, and ratings marked as missing were excluded prior to analysis.

For each clinician label, a bagged decision tree ensemble (200 trees, minimum leaf size of 10) was trained to predict the label from the three features. Model performance was estimated using repeated stratified 5-fold cross-validation with 5 repeats (25 folds total). Within each fold, features were standardized using the mean and standard deviation of the training partition only, and the same transformation was applied to the test partition to prevent data leakage.

Model discrimination was quantified differently depending on the number of classes. For the four binary labels, the area under the receiver operating characteristic curve (AUROC) was computed with respect to the positive class (e.g., “Present”), and precision, recall, and F1 score were reported for that class. For the three-class frequency label and the five-class diagnosis label, a one-vs-rest macro-averaged AUROC was computed across all classes per fold, and macro-averaged precision, recall, and F1 were calculated by averaging class-wise metrics, treating each class symmetrically. For the diagnosis label, micro-averaged F1 was additionally reported to account for class imbalance. All metrics were averaged across folds and repeats, with standard deviations reported.

Feature importance was assessed using permutation importance: for each feature, its values in the test partition were randomly shuffled — 10 times per fold for binary and frequency labels, and 30 times per fold for the diagnosis label to improve stability given the larger number of classes — and the resulting drop in AUROC relative to the baseline was recorded. For binary labels this used the standard AUROC drop; for the frequency and diagnosis labels the drop was computed on the macro-averaged one-vs-rest AUROC. Drops were averaged across all folds and repeats, clamped at zero to remove numerical noise, and normalized to sum to 100% to yield a percent

## Results

Four raters evaluated 102 tremor recordings, evenly distributed across rest, postural, and kinetic tasks from 34 participants. Interrater agreement on TWMQ items varied by feature (Table 1). Observed agreement ranged from 51% (Amplitude) to 79% (Tremor presence). Chance-corrected reliability (AC1/AC2) was substantial for Tremor presence (AC1 = 0.65) and Symmetry (three categories; AC1 = 0.69) despite marked class imbalance for Symmetry (asymmetric = 85%). Frequency (ordered, three levels) showed moderate agreement (AC2 = 0.47) with a predominance of the “mid” category (74%). Jerky and Rhythm demonstrated fair agreement (AC1 = 0.34 and 0.32, respectively), whereas amplitude showed slight agreement (AC1 = 0.014) despite balanced classes (constant = 52%, varies = 48%). The lowest level of agreement was observed for tremor diagnosis, with 32% observed agreement and a low reliability score (AC1 = 0.16).

**Table 1:**
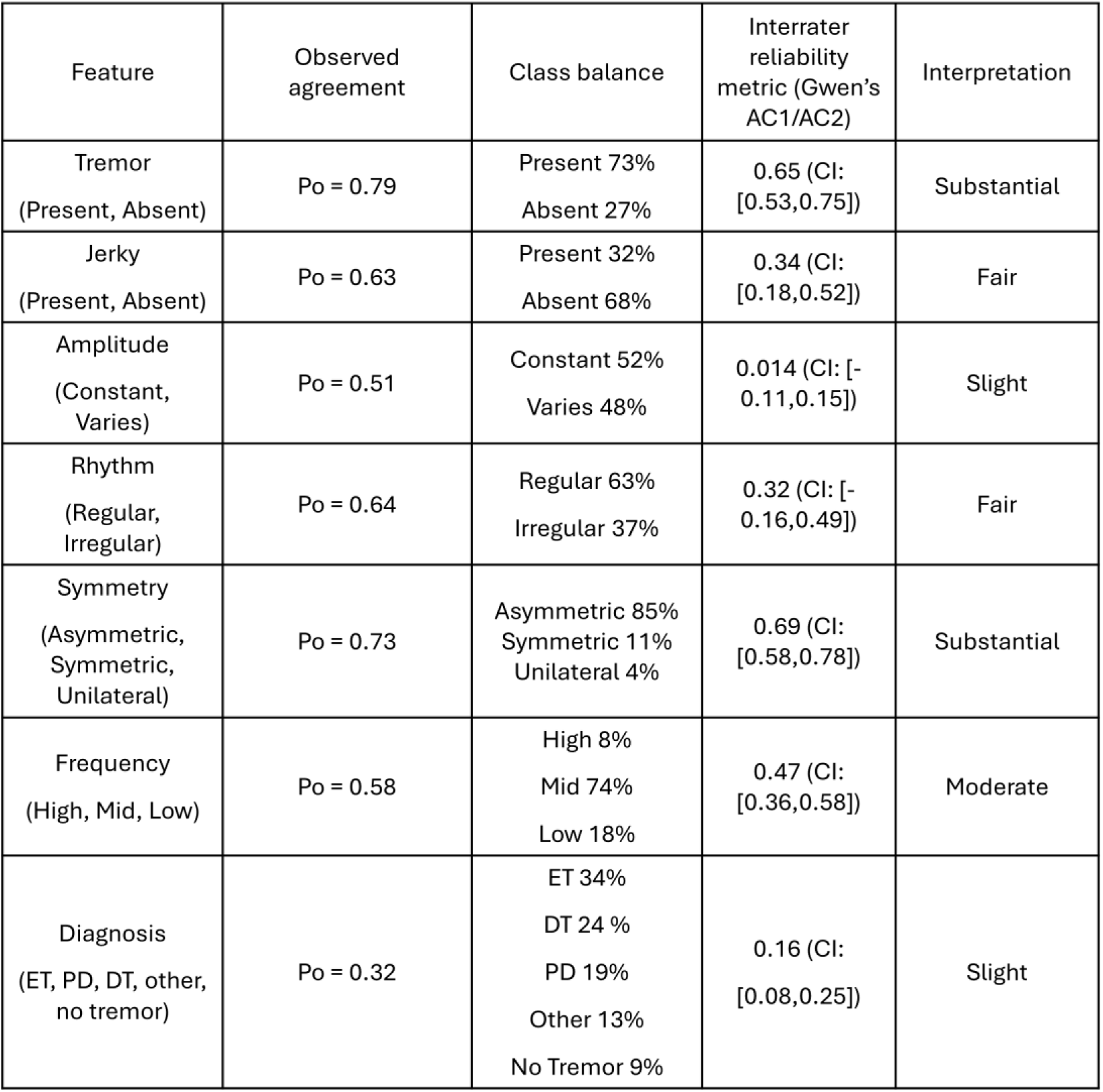
Interrater agreement for TWMQ items: Columns show Observed agreement (%), Class balance (per-category counts and percentages across all targets and raters), the interrater reliability metric (AC1 for nominal items; AC2 with quadratic weights for the ordinal frequency item), and a qualitative interpretation (“slight,” “fair,” “moderate,” “substantial,” “almost perfect”). CIs are 95% and were computed [analytic/by cluster bootstrap over targets]. *Interpretation guide (conventional, for readability):* slight <0.20, fair 0.21–0.40, moderate 0.41–0.60, substantial 0.61–0.80, almost perfect >0.80 as per Walsh et al. (15)

We assessed cross-feature effects to test whether classifying one attribute biased judgments of another. Table 2 reports the percentage distribution of ratings for amplitude × jerkiness, amplitude × rhythm, and rhythm × jerkiness, with cell entries indicating how often each combination occurred. All three pairs showed significant associations by χ², with effect sizes from moderate to large. The strongest association was rhythm × jerkiness (χ² = 154.38, *p* < 0.001, φ = 0.731; large), followed by amplitude × rhythm (χ² = 88.71, *p* < 0.001, φ = 0.556; large) and amplitude × jerkiness (χ² = 51.68, *p* < 0.001, φ = 0.425; moderate–large). Marginally, amplitude was classified as constant and varying in approximately equal proportions. Conditionally, when amplitude was constant, tremor was labeled non-jerky in >85% of ratings; when amplitude varied, jerky and non-jerky labels occurred with roughly equal frequency, and when jerkiness was present, ∼78% of ratings assigned varying amplitude. Likewise, with constant amplitude, >85% of ratings categorized rhythm as regular; when amplitude varied, rhythm was more often irregular (∼65%) than regular (∼35%), and irregular rhythm coincided with variable amplitude in ∼84% of ratings. For rhythm × jerkiness, regular rhythm corresponded to no jerkiness in 93% of ratings, whereas irregular rhythm corresponded to jerkiness in 76%. Together, these patterns indicate strong coupling between irregular rhythm and jerkiness, and between regular rhythm and non-jerky classifications, with amplitude variability aligning with the latter pairwise relationships (Table 2).

**Table 2:** Combined cross-feature comparisons: On the left side, each matrix summarizes joint classifications across all rater observations: (i) Jerkiness (present/absent) × Amplitude stability (constant/varying), (ii) Amplitude stability (constant/varying) × Rhythmicity (regular/irregular), and (iii) Rhythmicity (regular/irregular) × Jerkiness (present/absent). Cell entries are percentages computed after pooling ratings across rates. Missing entries were excluded for these calculations. This pooled view highlights co-occurrence patterns (e.g., irregular rhythm with varying amplitude or jerkiness). The right side shows the conditional probabilities for the corresponding comparisons (e.g. P_Jerky|Amplitude_ and P_Amplitude|Jerky_ for the (Amplitude vs Jerkiness) comparison.

| Cross-feature probabilities |  |  | Conditional Probabilities |  |  |  |  |  |
| --- | --- | --- | --- | --- | --- | --- | --- | --- |
| Amplitude Vs Jerkiness |  |  |  |  |  |  |  |  |
| Jerky Rating<br>Amplitude | Absent (%) | Present (%) | $P_{\text{Jerky}\vert \text{Amplitude}}$ | Absent (%) | Present (%) | $P_{\text{Amplitude}\vert \text{Rhythm}}$ | Absent (%) | Present (%) |
| Constant | 45.11 | 6.99 | Constant | 86.58 | 13.42 | Constant | 66.84 | 21.51 |
| Varies | 22.38 | 25.52 | Varies | 46.72 | 53.28 | Varies | 33.16 | 78.49 |
| Amplitude Vs Rhythm Rating |  |  |  |  |  |  |  |  |
| Rhythm Rating<br>Amplitude | Regular (%) | Irregular (%) | $P_{\text{Rhythm}\vert \text{Amplitude}}$ | Regular (%) | Irregular (%) | $P_{\text{Amplitude}\vert \text{Rhythm}}$ | Regular (%) | Irregular (%) |
| Constant | 45.99 | 5.92 | Constant | 88.59 | 11.41 | Constant | 73.33 | 15.89 |
| Varies | 16.73 | 31.36 | Varies | 34.78 | 65.22 | Varies | 26.67 | 84.11 |
| Rhythm Rating Vs Jerkiness |  |  |  |  |  |  |  |  |
| Jerky Rating<br>Rhythm Rating | Absent (%) | Present (%) | $P_{\text{Jerky}\vert \text{Rhythm}}$ | Absent (%) | Present (%) | $P_{\text{Rhythm}\vert \text{Jerky}}$ | Absent (%) | Present (%) |
| Regular | 58.82 | 3.81 | Regular | 93.92 | 6.08 | Regular | 87.18 | 11.70 |
| Irregular | 8.65 | 28.72 | Irregular | 23.15 | 76.85 | Irregular | 12.82 | 88.30 |

Using the parameters of peak amplitude, peak frequency, and frequency variability, we predicted six clinician-assigned tremor labels using a bagged ensemble classifier with repeated stratified 5-fold cross-validation. The percent influence of each feature on classification performance for all six labels is shown in Figure 1. Tremor presence was the best-discriminated label (Figure 1A; accuracy = 0.740, F1 = 0.839, AUROC = 0.749, 95% CI [0.727–0.771]), with peak amplitude emerging as the dominant feature (42.9%), followed by frequency variability (33.4%) and peak frequency (23.6%). Rhythmicity (Figure 1C; accuracy = 0.631, F1 = 0.728, AUROC = 0.673, 95% CI [0.650–0.697]) was driven predominantly by frequency variability (66.0%), with secondary contributions from peak amplitude (23.7%) and peak frequency (10.4%). Jerkiness (Figure 1D; accuracy = 0.683, F1 = 0.297, AUROC = 0.600, 95% CI [0.574–0.625]) was most strongly influenced by peak amplitude (55.4%), with meaningful contributions from frequency variability (38.2%) and a smaller effect of peak frequency (6.4%), though the low F1 relative to accuracy suggests a class imbalance favoring the absent category. Amplitude consistency (Figure 1B; accuracy = 0.517, F1 = 0.470, AUROC = 0.506, 95% CI [0.472–0.541]) performed near chance overall, yet was most influenced by frequency variability (54.9%), followed by peak frequency (27.9%) and peak amplitude (17.2%), suggesting that while these features carry some relevant information, they are insufficient to reliably discriminate this perceptual quality. Perceived tremor frequency (Figure 1E; accuracy = 0.731, Macro-F1 = 0.286, Macro-AUROC = 0.471, 95% CI [0.444–0.498]) was, as expected, dominated by peak frequency (62.4%), with smaller contributions from frequency variability (24.1%) and peak amplitude (13.5%), though the near-chance AUROC alongside a relatively high accuracy suggests the model defaulted toward the majority class. Clinical diagnosis (Figure 1F; accuracy = 0.318, Macro-F1 = 0.191, Macro-AUROC = 0.507, 95% CI [0.490–0.523]) performed near chance across all five categories, with peak amplitude as the most influential feature (62.1%), followed by peak frequency (24.3%) and frequency variability (13.6%), suggesting that diagnosis depends on signal characteristics beyond those captured by the three features examined here. To account for substantial interrater disagreement in diagnosis, we repeated the analysis using only cases with a consensus diagnosis (≥3 of 5 raters in agreement). Although overall diagnostic performance remained poor (data not shown), the relative importance of the objective features was similar, with peak amplitude contributing 52.0%, peak frequency 32.7%, and frequency variability 14.7%. These findings suggest that while objective tremor characteristics influence diagnostic impressions, clinical diagnosis relies on additional features not captured by the three-signal metrics examined here.

**Figure 1.**
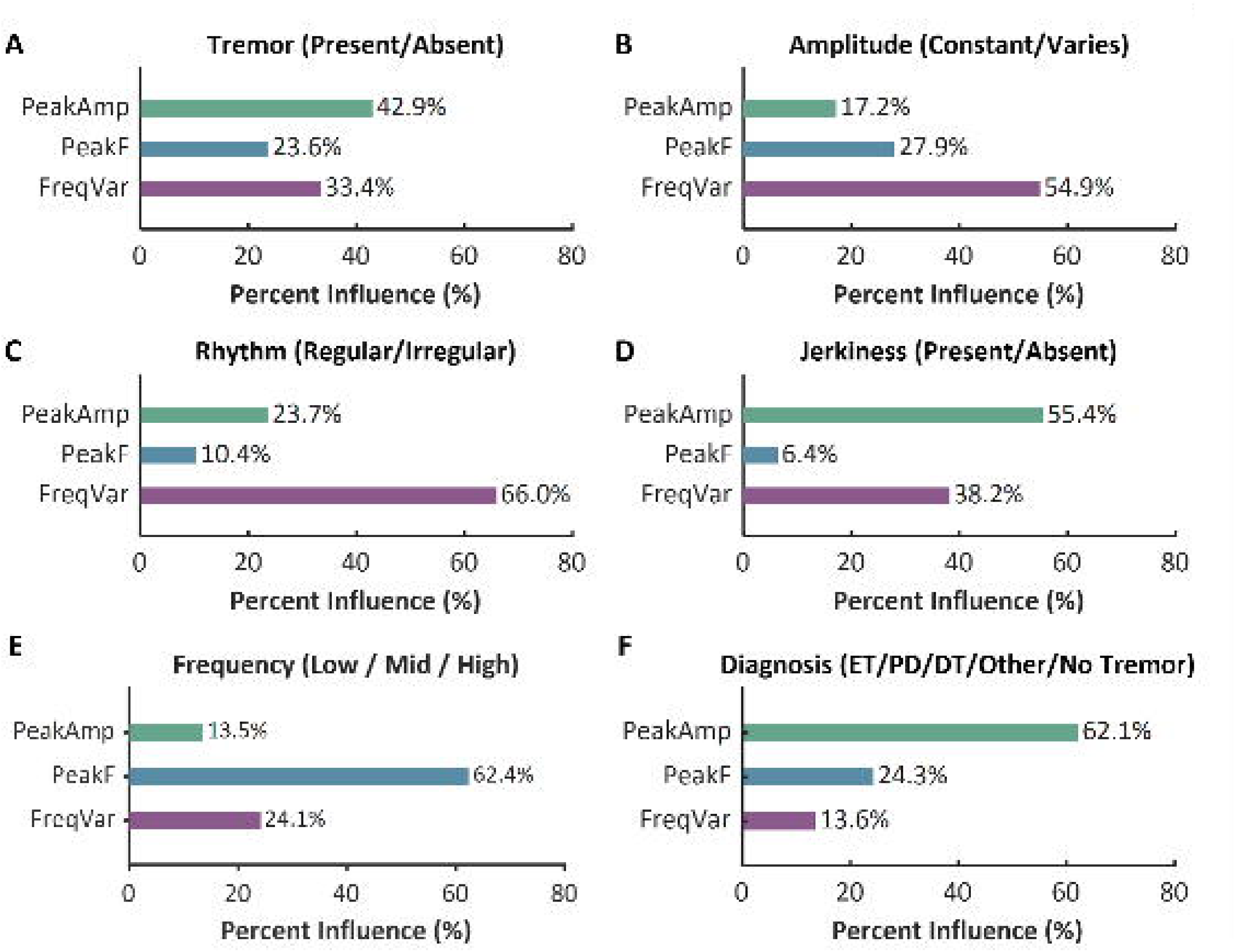
Feature dependence of clinician labels: Percent influence (permutation-based) of each objective feature on classification performance for: (A) Tremor (Present/Absent), (B) Amplitude (Constant/Varies), (C) Rhythm (Regular/Irregular), (D) Jerky (Present/Absent), (E) Frequency (Low/Mid/High), and (F) Diagnosis.

## Discussion

Using our developed TWMQ, we investigated the degree to which movement disorder neurologists consistently interpret key aspects of tremor morphology, including shape, rhythm, and regularity. These descriptors are widely employed in clinical assessments, published literature, and diagnostic reasoning. Overall, inter-rater agreement was relatively high for fundamental judgments, such as tremor presence and symmetry, but substantially lower for more nuanced descriptors including waveform shape, rhythmicity, and consistency of amplitude. This variability in rating consistency likely reflects a combination of factors: insufficiently defined or overlapping constructs, heterogeneity in raters’ conceptual interpretation of terminology, the inherent subtlety of the perceptual signs being evaluated, and the patterned co-occurrence of tremor features that may introduce systematic bias in clinical judgment.

### Tremor: Presence, Symmetry, Frequency

Among the questionnaire features, agreement was strongest for tremor presence, suggesting that clinicians can identify tremor with substantial reliability. However, even this relatively straightforward item showed only 0.76 observed agreement. This may reflect variability in how clinicians define or distinguish tremor from dystonia. Alternatively, it may reflect limitations of video-based assessment and highlight the value of objective measures such as accelerometry. Symmetry also demonstrated substantial interrater agreement. Despite a strong skew toward “asymmetric” (85%), the AC1 remained substantial—reflecting its robustness to marginal imbalances—and clinicians reliably classified lateralization as predominantly asymmetric, symmetric, or unilateral. Frequency showed moderate reliability, suggesting that ordinal distinctions among low/mid/high are usable but would benefit from tighter anchoring. For frequency, the dominance of “mid” (74%) hints at a default category in borderline cases; more precise anchors (e.g., low < 4 Hz, mid = 4–7 Hz, high > 7 Hz) or brief calibration modules that count cycles over fixed windows could mitigate this central-tendency bias.

### Tremor: Jerkiness, Rhythm, Amplitude

In contrast, cycle-to-cycle features showed lower interrater reliability. Jerkiness and rhythm achieved only fair agreement. This pattern likely reflects construct and/or rubric ambiguity rather than prevalence effects: jerkiness and rhythm overlap conceptually—cycle-to-cycle irregularity can present as both. The “constant vs. varies” distinction may lack operational thresholds when judged visually from video. Amplitude stability showed “slight” reliability despite near-balanced classes. Although amplitude is commonly assessed, the subtle cycle-to-cycle variability captured here did not yield strong agreement, suggesting rater subjectivity rather than a statistical artifact. In practice, all tremors exhibit some variability; accordingly, the threshold for labeling amplitude as “variable” versus “constant” appears to differ substantially across raters.

### Cross-feature comparisons of cycle morphology

Cross-feature analyses revealed systematic coupling among morphology descriptors. The strongest association was between rhythm and jerkiness, followed by amplitude–rhythm and amplitude–jerkiness. Directionally, constant amplitude co-occurred with regular rhythm and non-jerky labels, whereas varying amplitude co-occurred with irregular rhythm and jerkiness. Collectively, these patterns indicate that raters’ labels for “jerkiness,” “irregular rhythm,” and “variable amplitude” often index a common latent dimension of cycle-to-cycle irregularity rather than three distinct constructs. This overlap may reflect genuine co-occurrence in the underlying signal, how raters naturally perceive related movement patterns, or the traditional use of terms such as “jerky” and “irregular” to describe certain tremor types, which may lead raters to assign these labels together.

### Clinician’s perception and objective measure

Modeling of rater responses revealed that distinct perceptual judgments of tremor characteristics are differentially informed by underlying signal features. Tremor presence was primarily determined by amplitude, consistent with the intuitive notion that stronger oscillatory magnitude makes tremor more perceptually salient. However, both frequency variability and peak frequency also influenced this judgment, suggesting that raters implicitly integrate temporal stability when deciding whether a tremor is present. Amplitude classification (constant versus varying) and rhythm (regular versus irregular) were both driven predominantly by frequency variability, indicating that raters rely heavily on temporal fluctuations to distinguish stable from unstable tremor patterns. However, the temporal fluctuations and fluctuations in amplitude may have separate neurophysiological driver. These results suggest the interchangeability of perception of amplitude irregularity and irregular rhythm similar to discussed in cross-feature comparisons section. Judgments of jerkiness, in contrast, depend highly on the amplitude, this may occur as the amplitude increases, the jerky behavior may become more apparent to the rater. The models perform poorly for amplitude variability and jerkiness, as reflected by their low accuracy and AUC values. This likely indicates two possibilities. First, substantial variability or inconsistency in the labels may be limiting the model’s ability to learn reliable patterns. Second, the current features may not adequately capture the underlying characteristics of amplitude variability and jerkiness, highlighting the need to identify or derive a more appropriate set of parameters for these phenomena. Finally, frequency estimation tracked the objective peak frequency closely, yet the TWMQ showed only 58% agreement for the frequency item, implying that bin boundaries (e.g., low–medium, medium–high) introduce subjectivity near cut points.

### Tremor: Diagnosis

Considering the substantial disagreement observed across individual tremor features, the reliability of tremor diagnosis, which is the goal for patient care, is also compromised. Tremor diagnosis showed the lowest interrater agreement among all evaluated items. The current dataset suggests that tremor amplitude is a major factor influencing clinician perception when classifying tremor into categories such as ET, PD, Tremor with Dystonia, or other tremor types. Due to inconsistent diagnostic labels across raters, the classifier’s performance was negatively affected and resulted in the lowest accuracy among all features. These findings suggest that the clinical features assessed in this study alone may not fully capture the information clinicians use when assigning a tremor diagnosis, and that additional features, such as bradykinesia, hypomimia, or position dependence, may also contribute to clinical decision-making.

Interestingly, the factors associated with tremor diagnosis differed depending on the analytical approach. When analyses were restricted to the most clinically consistent diagnoses (i.e., cases with greater rater agreement), tremor frequency emerged as the strongest contributor to diagnostic classification. In contrast, when individual clinician ratings were analyzed separately, tremor amplitude was the dominant factor influencing diagnostic decisions. These findings suggest that while frequency may be the primary feature underlying consensus among clinicians, amplitude may play a larger role in individual clinician perception when classifying tremor into categories such as ET, PD, DT, or other tremor types.

### Physiological correlations of cycle morphology

“Rhythmicity” and “regularity” are widely used yet loosely defined descriptors of tremor. No consensus scale specifies how much irregularity is compatible with labeling a movement as tremor, and in practice no tremor is perfectly periodic rhythmicity varies across conditions (11). Despite this vagueness, cycle-to-cycle variability likely encodes meaningful physiology. For instance, reduced amplitude has been linked to increased frequency variability, consistent with central oscillators drifting out of synchrony (15), while amplitude fluctuations themselves may reflect variability in the firing of individual neurons within the tremor-generating network (16).The findings underscore that analyzing signal structure/variability is crucial for discriminating against tremor forms when mean amplitude is indistinguishable. Morphology labels can also mislead “dystonic tremor” is often called “jerky” or “irregular” (12) and our cross-feature analysis showed raters frequently apply these labels together, yet prior work indicates dystonic with tremor can exhibit a regular rhythm (11,17) suggests that tremor mixed with dystonia can this type of tremor can have a regular rhythm. These observations underscore the need for operational definitions and quantitative anchors for rhythmicity and regularity.

### Implications for research and technological development

Literature frequently treats these waveform descriptors as clinically meaningful, yet formal, objective measurement standards are lacking. This gap matters: the same descriptors are used routinely in clinic and often serve as “ground truth” in studies training machine-learning classifiers to differentiate diagnoses (4,18–21). With the rise of AI approaches, imprecise or inconsistently applied labels risk propagating bias and limiting model generalizability. Our findings highlight the need to refine these features, establish clear operational definitions, and integrate objective instrumentation (e.g., sensor-based metrics) to improve the reliability, interpretability, and translational value of tremor waveform characterization.

## Conclusion

Clinically, these findings support using presence and symmetry as dependable descriptors for communication, whereas morphology-level labels: jerkiness, rhythm, and amplitude stability should be interpreted with caution because they can overlap conceptually and risk misclassification. For research and diagnostic workflows, pairing rater judgments with objective signals to establish clear anchors can improve accuracy and consistency.

## Data Availability

All data produced in the present study are available upon reasonable request to the authors after appropriate approval processes.

## Author’s Roles

- PJ: Writing – original draft, data analysis, Visualization
- YB: Execution, Data Collection, Writing – review & editing
- SP: Investigation, Writing – review & editing
- MV: Investigation, Writing – review & editing
- VF: Writing – review & editing
- MH: Conceptualization, Investigation
- HJ: Investigation, Writing – review & editing
- AGS: Conceptualization, Methodology, Investigation, Project administration, Funding acquisition, Supervision, Writing – review & editing
- AF: Conceptualization, Methodology, Investigation, Project administration, Funding acquisition, Supervision, Writing – review & editing

